# What is required to prevent a second major outbreak of the novel coronavirus SARS-CoV-2 upon lifting the metropolitan-wide quarantine of Wuhan city, China

**DOI:** 10.1101/2020.03.24.20042374

**Authors:** Lei Zhang, Mingwang Shen, Xiaomeng Ma, Shu Su, Wenfeng Gong, Jing Wang, Yusha Tao, Zhuoru Zou, Rui Zhao, Joseph Lau, Wei Li, Feng Liu, Kai Ye, Youfa Wang, Guihua Zhuang, Christopher K Fairley

**Affiliations:** China-Australia Joint Research Center for Infectious Diseases, School of Public Health, Xi’an Jiaotong University Health Science Center, Xi’an, Shaanxi, 710061, PR China; Melbourne Sexual Health Centre, Alfred Health, Melbourne, Australia; Central Clinical School, Faculty of Medicine, Nursing and Health Sciences, Monash University, Melbourne, VIC, Australia; Department of Epidemiology and Biostatistics, College of Public Health, Zhengzhou University, Zhengzhou 450001, Henan, China; Institute of Health Policy Management and Evaluation, University of Toronto, Toronto, Ontario, Canada; School of Public Health and Preventive Medicine, Faculty of Medicine, Monash University, Melbourne, Australia; Centre for Research Behaviors, The Jockey Club School of Public Health and Primary Care, The Chinese University of Hong Kong, HKSAR, China; Bill and Melinda Gates Foundation, Seattle, USA; Department of Epidemiology and Health Statistics, School of Public Health, Southeast University, Nanjing, Jiangsu, China; Shaanxi Provincial Center for Disease Control and Prevention, Xi’an, 710054, Shaanxi, China; MOE Key Lab for Intelligent Networks & Networks Security, Faculty of Electronics and Information Engineering, Xi’an Jiaotong University, Xi’an, 710049 China; School of Automation Science and Engineering, Faculty of Electronics and Information Engineering, Xi’an Jiaotong University, Xi’an, 710049 China; Genome Institute, the First Affiliated Hospital of Xi’an Jiaotong University, Xi’an, 710061 China; The School of Life Science and Technology, Xi’an Jiaotong University, Xi’an, 710049 China; Global Health Institute, Department of Epidemiology and Biostatistics, School of Public Health, Xi’an Jiaotong University Health Science Center, Xi’an, Shaanxi, 710061, China

**Keywords:** COVID-19, epidemic projection, metropolitan-wide quarantine, mathematical modelling

## Abstract

**Background:** The Chinese government implemented a metropolitan-wide quarantine of Wuhan city on 23rd January 2020 to curb the epidemic of the coronavirus COVID-19. Lifting of this quarantine is imminent. We modelled the effects of two key health interventions on the epidemic when the quarantine is lifted.

**Method:** We constructed a compartmental dynamic model to forecast the trend of the COVID-19 epidemic at different quarantine lifting dates and investigated the impact of different rates of public contact and facial mask usage on the epidemic.

**Results:** We estimated that at the end of the epidemic, a total of 65,572 (46,156-95,264) individuals would be infected by the virus, among which 16,144 (14,422-23,447, 24.6%) would be infected through public contacts, 45,795 (32,390-66,395, 69.7%) through household contact, 3,633 (2,344-5,865, 5.5%) through hospital contacts (including 783 (553-1,134) non-COVID-19 patients and 2,850 (1,801-4,981) medical staff members). A total of 3,262 (1,592-6,470) would die of COVID-19 related pneumonia in Wuhan. For an early lifting date (21st March), facial mask needed to be sustained at a relatively high rate (≥85%) if public contacts were to recover to 100% of the pre-quarantine level. In contrast, lifting the quarantine on 18th April allowed public person-to-person contact adjusted back to the pre-quarantine level with a substantially lower level of facial mask usage (75%). However, a low facial mask usage (<50%) combined with an increased public contact (>100%) would always lead a significant second outbreak in most quarantine lifting scenarios. Lifting the quarantine on 25th April would ensure a smooth decline of the epidemics regardless of the combinations of public contact rates and facial mask usage.

**Conclusion:** The prevention of a second epidemic is viable after the metropolitan-wide quarantine is lifted but requires a sustaining high facial mask usage and a low public contact rate.

---

The outbreak of the novel coronavirus SARS-CoV-2 was first identified in the Chinese city of Wuhan in early December 2019, when a group of 27 patients with close contact with a seafood market were diagnosed with a pneumonia of unknown aetiology ^1,2^. The virus was found to be highly contagious and transmit in populations via droplet, person-to-person contact and aerosol transmission ^3,4^. The number of infected cases increased rapidly in Wuhan during the first few weeks of the outbreak, and then quickly spread to all 31 Chinese provinces and aboard ^5,6^. As of 20^th^ March 2020, 180 countries worldwide have reported cases of COVID-19. So far, China has reported 80,695 confirmed cases and 3,097 deaths, accounting for about one-third of all cases and deaths worldwide.

To curb the epidemic, the Chinese government introduced a ‘metropolitan-wide quarantine’ of the city of Wuhan from 23^rd^ January 2020, by terminating all public transportation in the city and intercity links ^7^. During the metropolitan-wide quarantine, the National Health Commission and the China Centre for Disease Control urged the use of facial masks in all public spaces, put in place strict home containment policies, postponed schools and industry reopening to reduce communal activities and person-to-person transmission. A massive screening program was implemented for individuals in close contact with the infected or high-risk individuals ^8-10^. The strict control in Wuhan has been effective, with the daily reported confirmed cases significantly reduced from 1500-2000 at its peak to 10 cases or less a day^11^. However, the implementation of the quarantine has also severely damaged its economy, with predictions that the Chinese economy will grow by less than 4% in the first quarter of 2020 ^12^. The daily life of Wuhan residents has also been seriously disrupted, and the long duration of home containment may result in mental and psychological issues ^13,14^.

Lifting the metropolitan-wide quarantine is imminent. Since late February, major cities across China have gradually eased their restriction levels and partially resumed public transportation^15^. As the epicentre of the outbreak, Wuhan faces the dilemma of balancing the substantial accumulating economic losses with the hard-earned control of the epidemic. Lifting the quarantine restrictions in the city and reopening transport links with the rest of China has become the top priority for the policymakers. We aim to determine to what level of the two commonly used control measures, social distancing and facial mask usage, are necessary to prevent a resurgence of the epidemic due to either residual active cases in Wuhan or imported cases after lifting the quarantine.

We estimated that at the end of the epidemic, a total of 65,572 (46,156-95,264) individuals would be infected by the virus, among which 16,144 (14,422-23,447, 24.6%) would be infected through public contacts, 45,795 (32,390-66,395, 69.7%) through household contact, 3,633 (2,344-5,865, 5.5%) through hospital contacts (including 783 (553-1,134) non-COVID-19 patients and 2,850 (1,801-4,981) medical staff members). A total of 3,262 (1,592-6,470) would die of COVID-19 related pneumonia in Wuhan.

We examined six proposed dates for quarantine lifting in our model. When public contact recovered to 100% the pre-quarantine level and facial mask usage was high at 95%, the epidemic would follow a smooth decline to elimination regardless of which day for quarantine was lifted (Figure 1a). In contrast, when facial mask usage was reduced to 50%, any quarantine lifting date before 25^th^ April would result in a second major outbreak (Figure 1b). Similarly, when facial mask usage was sustained at 80%, and public contact rate was recovered to 100% of the pre-quarantine level, an earlier lifting on 21^st^ March may lead to a second minor outbreak (Figure 1c). But, if the public contact rate was 50% more than the pre-quarantine level, a second major outbreak would occur in all quarantine lifting dates except 25^th^ April (Figure 1d).

**Figure 1.**
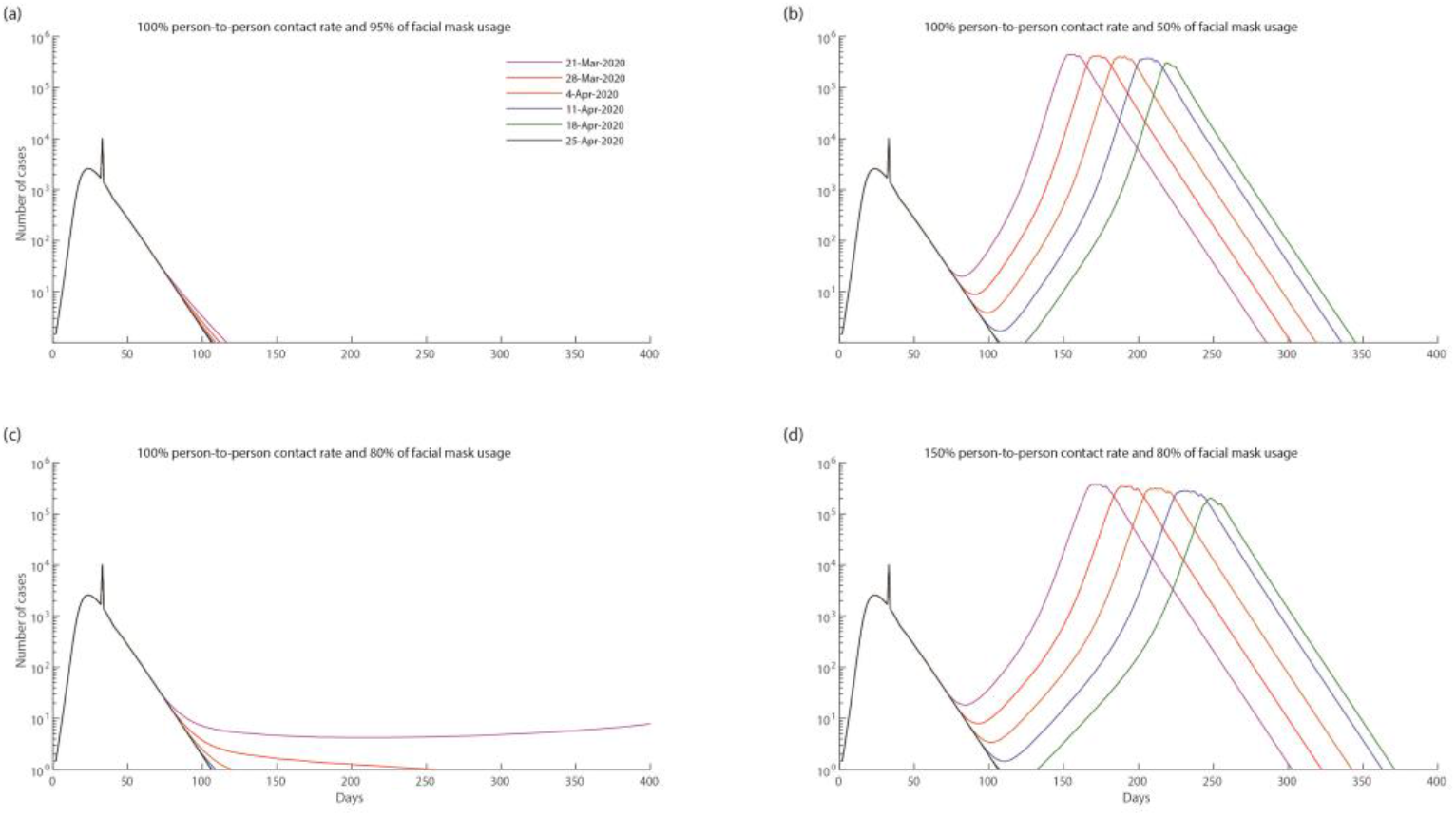
a-b Impact of different levels of facial mask usage (95% and 50%) on the COVID-19 epidemic in Wuhan city, on various quarantine lifting dates. The public contact rate is fixed at 100% (same as the pre-quarantine level). c-d Impact of different levels of public contact rate (80% and 150%) on the COVID-19 epidemic in Wuhan city, on various quarantine lifting dates. The facial mask usage is reduced to 80%.

Combinations of high facial mask usage and reduced public contacts may lead to a smooth decline of the epidemic on various quarantine lifting dates (Figure 2). For an early lifting date (21^st^ March), facial mask needed to be sustained at a relatively high rate (≥85%) if public contacts were to recover to 100% of the pre-quarantine level. In contrast, lifting the quarantine on 18^th^ April allowed public person-to-person contact adjusted back to the pre-quarantine level with a substantially lower level of facial mask usage (75%). However, a low facial mask usage (<50%) combined with an increased public contact (>100%) would always lead a significant second outbreak in most quarantine lifting scenarios. Lifting the quarantine on 25^th^ April would ensure a smooth decline of the epidemics regardless of the combinations of public contact rates and facial mask usage.

**Figure 2.**
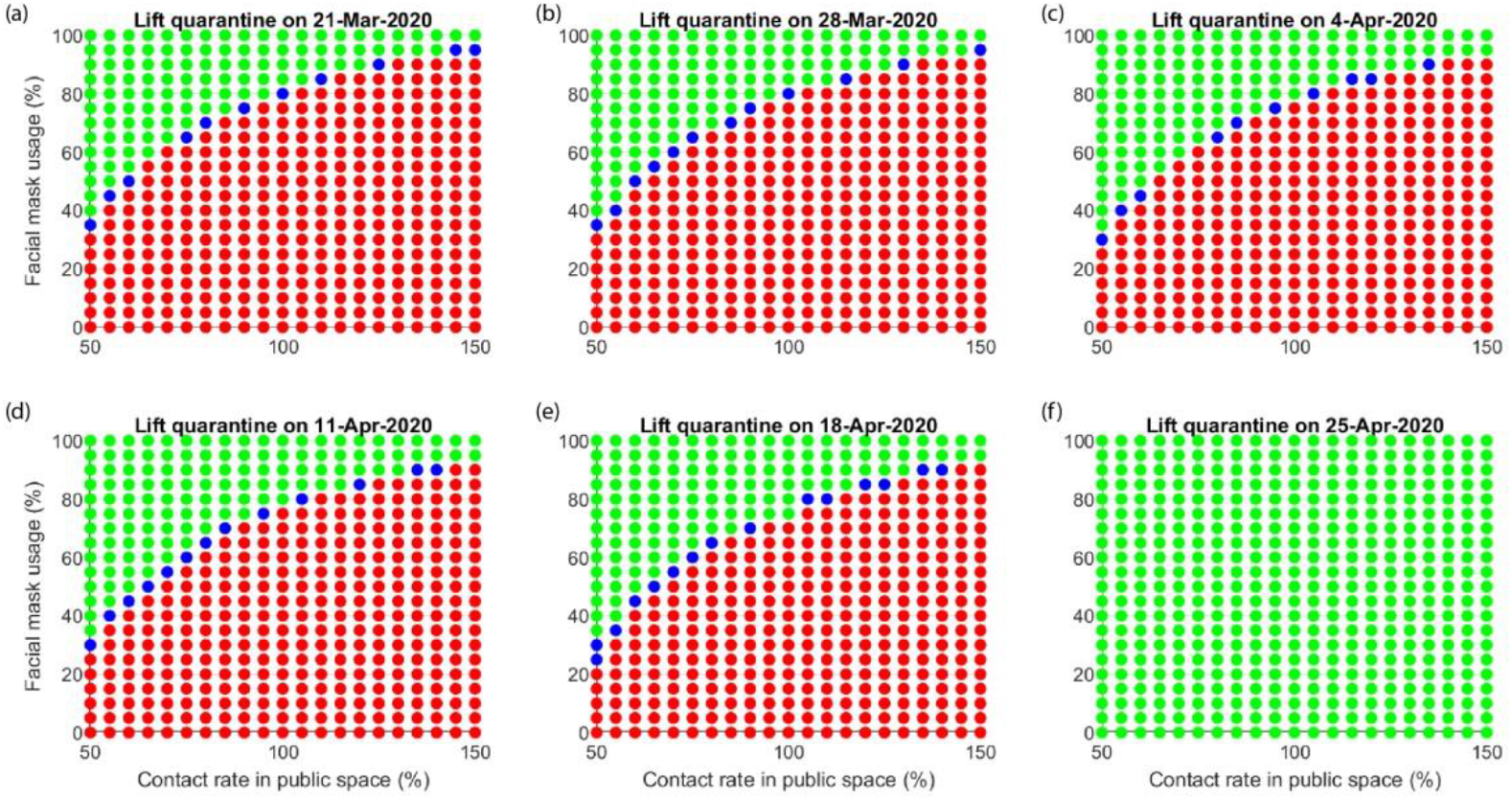
Impact of various combination of facial mask usage and public person-to-person contact rate on the COVID-19, on various quarantine lifting dates including hypothetically (f) waiting till all internal cases resolved and continuing a travel ban. (Green: smooth declining epidemic after lifting; Blue: a second minor outbreak that is less than the current outbreak; Red; a second major outbreak that is greater than the current outbreak)

Metropolitan-wide quarantine that was introduced in Wuhan on 23rd January 2020 and has been remarkably effective at reducing transmission of COVID-19. We showed that it was possible to lift the quarantine and control the epidemic, but only if specific measures such as sustaining high facial mask usage and limiting the public contact rate remained in place. Otherwise, the epidemic will recur either from residual cases in Wuhan or imported cases from elsewhere when the transport links resume. Adopting and maximising other methods such as testing, contact tracing, and frequent hand washing would also reduce the probability of a second epidemic.

The quarantine has substantially altered the transmission pattern of the virus in Wuhan. While our model predicts that public transmission accounts for the majority of transmission prior to the quarantine, household transmission is the dominant route of transmission during the quarantine. This is intuitively reasonable as the quarantine has significantly reduced the public contacts of the residents and increased their contacts with family members in a closed household environment. In cases where the epidemic is able to be contained without a second outbreak, the dominant route of transmission would switch back to the public transmission after lifting the quarantine (Figure S4a). In cases where the epidemic results in a second major outbreak (Figure 4b), the second outbreak would be predominantly driven by household transmission. Our estimate that ∼70% of all infected cases was due to household transmission is also in a broad park agreement with a recent report of WHO (78-85% ^16^). Further, we estimated 5% hospital-acquired infections in (patients, 1.1%; medical staff 3.8%), which is consistent with a recent clinical report ^17^.

Our study suggests that maintaining a reduction in the contact rate below the pre-quarantine levels is an important ongoing intervention until a vaccine is available. If residents return to the same level of activities as the pre-quarantine very high level of facial mask usage rate of 85% (or other powerful interventions) will be required. Any additional public interactions, such as the influx of the five million returning residents to Wuhan, may trigger a second outbreak. Governments will need to determine how to minimise public contacts from workplaces, venues for essential daily commodities, residents, leisure, entertainment venues and public events although limiting or modifying the later may be important. Intercity travel should be minimised, and the return of the residents may need to be staged. The full restoration of the intercity public transportation may take months, and careful planning of the size of population inflow is necessary.

Maintaining ongoing high facial mask usage among the population may be challenging for a number of reasons. Firstly, supplying this number of masks to a population of the size of Wuhan let alone other cities in China and the world will be very challenging and may critically limit their availability for health care workers at high risk. Re-using disposable masks that may limit their efficacy.

Our findings need to be interpreted with caution. Our predictions are based on an assumption of a homogenous population, and in reality, human behaviours and interactions are far heterogeneous and complex. Population-dense areas are prone to virus transmission and sporadic outbreaks. High-risk hotspots are needed to be identified ahead and subjected to further monitoring after quarantine lifting. The decision regarding the degree to which the quarantine is lifted requires a balanced consideration of the direct economic cost of the quarantine. Given the current Chinese per-capita GDP is US$10,099, every day of the quarantine of Wuhan, a city of the population of over 10 million, means a direct economic loss of US$280m. The degree to which it is lifted should balance the economic implications and likelihood of a second outbreak. Further, the effect of handwashing was not quantified in this model due to very little field data informing this.

China is in a unique position to determine if the quarantine measures that successfully contained COVID-19 in Wuhan can be eased while allowing economic activity to resume. We recommend population facial mask usage only when the provision for high-risk health care workers is secured. We acknowledge the substantial pragmatic issues exist in rolling out and maintaining such a program worldwide due to inadequate supply of facial masks. But notwithstanding this limitation, this information may be of use to other cities, in other parts of Asia, Europe and the US that are currently experiencing rising epidemics of COVID-19. Limiting transmission while maintaining economic activity until a vaccine is available would be the ultimate goal.

## Data Availability

Data are given in attached Appendix

## Contributors

LZ and MS conceived the study. LZ, CKF contributed to the collection and interpretation of data. LZ, MS and JW did the data cleaning and statistical analysis. LZ conducted model building with assistance from MS. LZ, MS, XM and SS drafted the manuscript. All authors contributed to critical revision of the manuscript for important intellectual content. WG JW YT ZZ RZ JL WL FL KY YW GZ nad CKF provided epidemiological, technical, or material support. LZ and MS had full access to all the data in the study and take responsibility for the integrity of the data and the accuracy of the data analysis.

## Acknowledgments

This work is supported by a Research Grant from the Bill & Melinda Gates Foundation.

